# Short-Latency Changes in Closed-Eye EOG Following Medication Intake in ADHD

**DOI:** 10.1101/2025.11.23.25340824

**Authors:** Malthe Brændholt, Stefan Bjerrum

## Abstract

**Background:** Optimising medication for Attention Deficit Hyperactivity Disorder (ADHD) is often hindered by the lack of practical physiological indicators that can inform treatment effects. Electrooculography (EOG) has been suggested as a low-burden marker of neurophysiological processes relevant to ADHD, but its responsiveness to medication within routine clinical settings is not well established.

**Methods:** We analysed anonymised clinical records from adults with ADHD receiving stable, ongoing medication who underwent closed-eye EOG recordings immediately before and after taking their scheduled dose during standard consultations. Recordings were obtained using clinical EEG equipment. Heart rate and blood pressure, measured after each EOG session, were included as contextual indices of physiological state. Pre–post changes were assessed using paired t-tests and binomial tests, with Pearson correlations used to evaluate associations between EOG and cardiovascular measures.

**Results:** Both mean and peak EOG power showed consistent reductions following medication, with the majority of individuals exhibiting a decrease across measures. Cardiovascular parameters showed no systematic pre–post change, and differences in cardiovascular measures did not correlate with changes in EOG activity.

**Conclusions:** Closed-eye EOG showed systematic within-session modulation following medication intake in adults with ADHD on stable treatment. These findings provide preliminary support for EOG as a practical, non-invasive measure of short-latency medication-related neurophysiological effects, motivating further controlled studies to evaluate its potential relevance for treatment monitoring and titration.

## 1. Introduction

### 1.1 Clinical Challenge

Attention Deficit Hyperactivity Disorder (ADHD) is a complex neurodevelopmental disorder that significantly impacts individuals across their lifespan. The clinical burden associated with ADHD is substantial, with untreated cases leading to considerable economic costs, estimated at over €20,000 per adult annually due to increased healthcare utilisation, lower income, and higher reliance on state benefits compared to their non-ADHD counterparts (1) Furthermore, the challenges of medication titration complicate treatment adherence, as patients often experience difficulties in managing their treatment regimens, leading to non-compliance rates ranging from 13% to 64% (2) This non-adherence is exacerbated during the titration phase, where the need for careful dosage adjustments can discourage patients and their families, ultimately resulting in early discontinuation of treatment (3) The economic implications of delayed treatment due to titration challenges are significant, not only affecting the individual but also imposing a broader societal burden through increased healthcare costs and lost productivity (4) Addressing these issues is crucial for improving patient outcomes and optimising healthcare resources.

### 1.2 Personalised Medication In ADHD

The pursuit of personalised medication in ADHD is increasingly supported by emerging research on biomarkers. ADHD is characterised by considerable heterogeneity in symptom profiles and treatment responses, highlighting the need for approaches that accommodate individual variability. Several potential biomarkers have been identified that may help guide medication titration and improve treatment efficacy. For instance, genetic variants such as single nucleotide polymorphisms (SNPs) in genes including ADRA2A and COMT have been associated with differential responses to methylphenidate, suggesting a genetic basis for individual variation in treatment outcomes (5)

Neuroimaging and electrophysiological measures have also shown promise. Techniques such as electroencephalography (EEG) have revealed neural signatures that may predict stimulant response, offering a more targeted approach to treatment selection (6) Similarly, stress-related and immune markers measured in saliva have provided insight into physiological processes relevant to ADHD and may help track treatment effects over time (7)

Although these findings point to a growing toolbox of potential biomarkers, their practical value in routine clinical care remains to be established. A central question is whether such markers can deliver reliable, interpretable information that meaningfully supports medication titration in everyday settings. Demonstrating this will require robust validation, standardised protocols, and a clearer understanding of how biomarker signals interact with individual patient characteristics [(8) (9)]

### 1.3 EOG As A Neurophysiological Marker

Electrooculography (EOG) is a non-invasive technique that measures eye movements by detecting the cornea–retinal standing potential, providing low-burden, high-resolution physiological data well suited to settings where practicality and patient compliance are essential. EOG has been used to index a range of neurophysiological processes, including emotional responses and sleep stages, and has shown utility in state anxiety monitoring and sleep stage classification [(10) (11)] Altered EOG patterns have also been described across psychiatric conditions, reflecting underlying cognitive and emotional processes (12)

In ADHD, several studies indicate that EOG-derived features can distinguish affected individuals from healthy controls. These include eye movement behaviour in virtual reality tasks, which has demonstrated high classification accuracy (13) as well as diagnostic systems based on EOG alone (14) Differences in EOG signal characteristics, such as complexity measures, further support its relevance to ADHD-related neurophysiology (15)

Importantly, EOG remains informative in closed-eye conditions (11) Closed-eye movements can be reliably captured and show similarities to REM sleep eye movements in velocity and trajectory when visual input is absent [(16) (17)] This suggests that closed-eye recordings provide access to intrinsic oculomotor dynamics while minimising external sensory input.

Together, these findings highlight EOG as a practical and physiologically grounded method that can capture meaningful neurophysiological variation relevant to ADHD.

### 1.4 Hypothesis

Building upon prior evidence linking EOG alterations to ADHD traits, we hypothesise that medication may modulate intrinsic EOG activity in a manner consistent with changes in attentional regulation. Previous work indicates that EOG patterns can change transiently (10) suggesting a short-latency reactivity that may be informative within clinical timeframes. The continuity between REM and closed-eye awake eye movements further supports the use of closed-eye conditions as a stable, low-stimulus recording environment.

Therefore, we hypothesise that closed-eye EOG will be sensitive to medication-related neurophysiological changes within a single clinical session, providing a potential short-latency physiological window into medication related effects.

### 1.5 Study Aim

To test this hypothesis, we aim to evaluate the most direct prediction it generates within the constraints of the available clinical data: the EOG of patients with ADHD who receive optimal pharmacological treatment should show a consistent within-session change following medication intake and the intervening clinical procedures. Characterising this initial within-session relationship will allow future research to examine more complex aspects of EOG reactivity in relation to medication efficacy. If our hypothesis is not rejected, it will provide preliminary support for the idea that EOG responses may reflect medication-related neurophysiological changes, forming a basis for future controlled studies to assess their potential relevance for treatment evaluation.

## 2. Methods

### 2.1 Patient Data

This study was based exclusively on anonymised data derived from patient records. The original data were collected as part of standard clinical care for individuals treated for ADHD. As part of routine assessment and treatment, patients were informed about and given the option to undergo EOG recordings before and after medication, to be reviewed alongside other standard clinical measures such as blood pressure, heart rate, body mass index, clinical rating scales, and medication data.

Patients who accepted the option to undergo an EOG recording had been scheduled for their usual consultations in the mornings, with instruction not to take their regular morning medication until immediately after the first measurement. The clinical setting included a consultation room with comfortable seating, where two recording sessions of five minutes each were conducted with the patients’ eyes closed and awake. The procedure involved a premedication recording session followed by the administration of the medication, after which a normal consultation lasting approximately 30 minutes took place before the post-medication recording session.

For the present analysis, we included records from patients whose clinical data met the following criteria a diagnosis of ADHD in accordance with the DSM-5 (18) or ICD-10 (19), with a minimum DIVA score of six elements for inattention and/or six elements for hyperactivity/impulsivity; stable and satisfactory pharmacological treatment; and an age range of 18 to 55 years. Exclusion criteria comprised any diseases affecting eye movements and other significant psychiatric disorders.

### 2.2 EOG Acquisition

EOG data were recorded using standard clinical EEG equipment with a sampling frequency of 100 Hz. Electrodes were placed at the forehead and at the left and right epicanthus regions. The forehead electrode served as a reference for the two lateral electrodes.

### 2.3 Cardiovascular Measures

ADHD medications, such as dexmethylphenidate and guanfacine, have been shown to exert significant effects on cardiovascular function, including alterations in heart rate and blood pressure during both acute and long-term treatment phases (20) In addition, psychological and physical state is likely to vary between measurements. These potentially systematic autonomic and arousal-related changes could influence the EOG measures.

To address this potential source of bias, the blood pressure and heart rate measurements taken directly after each EOG session were interpreted as proxies for general arousal level and included as control parameters in the analysis. These measurements were obtained using a standard automated device.

### 2.4 Signal Processing

To quantify the frequency characteristics of the EOG signal, spectral analysis was conducted. The power spectral density was computed using Welch’s method, which employed a Hanning window with a segment length of 1024 samples. This approach facilitated a robust estimation of the power spectrum, allowing for the identification of significant frequency components within the EOG signal.

In this initial analysis, the EOG activity was parameterised by extracting key features from each recording of the patients. Specifically, the mean power and power at peak frequency were determined from the EOG power-frequency spectrum, forming the basis for further statistical analysis .

### 2.5 Statistical Analysis

Prior to statistical testing, both mean and peak EOG power values were normalised by dividing them by the corresponding median group-level pre-medication values. Initially, we conducted paired t-tests to assess whether the mean change from pre-to post-medication differed significantly from zero for mean and peak EOG power, as well as the cardiovascular measures, pulse, systolic, and diastolic blood pressure. This approach accounts for the paired nature of the data and quantifies the average magnitude and direction of change across participants, providing a robust measure of within-subject effects. In addition to the paired t-tests, we performed binomial (sign) tests to examine the directional consistency of the observed changes. This non-parametric test compares the proportion of participants exhibiting a consistent direction against a 50% chance level, allowing us to determine whether effects were systematically aligned across individuals. Together, these tests offer complementary insights into pre-to post-medication differences with the paired t-test focusing on mean magnitude and the binomial test assessing consistency across participants. To further explore the relationship between EOG and cardiovascular measures, we calculated Pearson correlations between individual differences (Δ = post-medication – pre-medication) in EOG and cardiovascular parameters. For each parameter pair, we reported correlation coefficients (r), two-tailed p-values, and the number of observations (n). All statistical tests were conducted at a standard alpha level of 0.05, which was used to guide the interpretation of statistical significance.

### 2.6 Analysis Code

The following Python packages were used for analysis: NumPy (21), SciPy (22), pandas (23), Matplotlib (24), Seaborn (25), MNE-Python (26), Pydantic (27).

## 3. Results

### 3.1 Participant Characteristics

In **Table 1**, we present the demographic and clinical characteristics of the participants involved in the study. A total of 20 candidates were included, with a mean age of 33years (SD = 9), and a median age of 32 years, ranging from 23 to 53 years. The participants’ scores on the Adult ADHD Self-Report Scale (ASRS) (28) at initial assessment averaged 56 (SD = 6), with a median score of 55, and scores ranging from 37 to 66. The change in ASRS scores (ASRS delta) indicated a mean decrease of -29 (SD = 7), with a median change of -29, and a range from -46.00 to -18.00. Gender distribution among participants revealed that 70% were female (n = 14) and 30% were male (n = 6) .

**Table 1:** Summary of participant characteristics. IQR: Interquartile range; SD; Standard deviation; ASR: Adult ADHD Self-Report Scale

| Variable | Statistic | Value |
| --- | --- | --- |
| **Age (years)** | Mean (SD) | 33 (9) |
|  | Median [IQR] | 32 [25–37] |
|  | Range | 23–53 |
| **Gender** | Female | 14 (70%) |
|  | Male | 6 (30%) |
| **ASRS (initial)** | Mean (SD) | 56 (6) |
|  | Median [IQR] | 55 [53–59] |
|  | Range | 37–66 |
| **ASRS ($\Delta$)** | Mean (SD) | −29 (7) |
|  | Median [IQR] | −28 [−31–24] |
|  | Range | −46–18 |

### 3.2 EOG Changes

In our analysis of EOG changes, we observed a consistent reduction in both mean power and peak power following the intervention. Specifically, for mean power, we found a significant decrease post-treatment, with a paired t-test yielding t(19) = -2.542, p = 0.020. Similarly, peak power also demonstrated a significant reduction, with t(19) = -2.331, p = 0.031. Furthermore, the binomial tests indicated that both EOG measures were aligned with this trend, as 18 out of 20 participants exhibited decreases in mean power and peak power, resulting in a binomial p-value of 0.0004 for both measures. These findings are illustrated in **Figure 1**, highlighting the substantial impact of the intervention on EOG activity.

**Figure 1:**
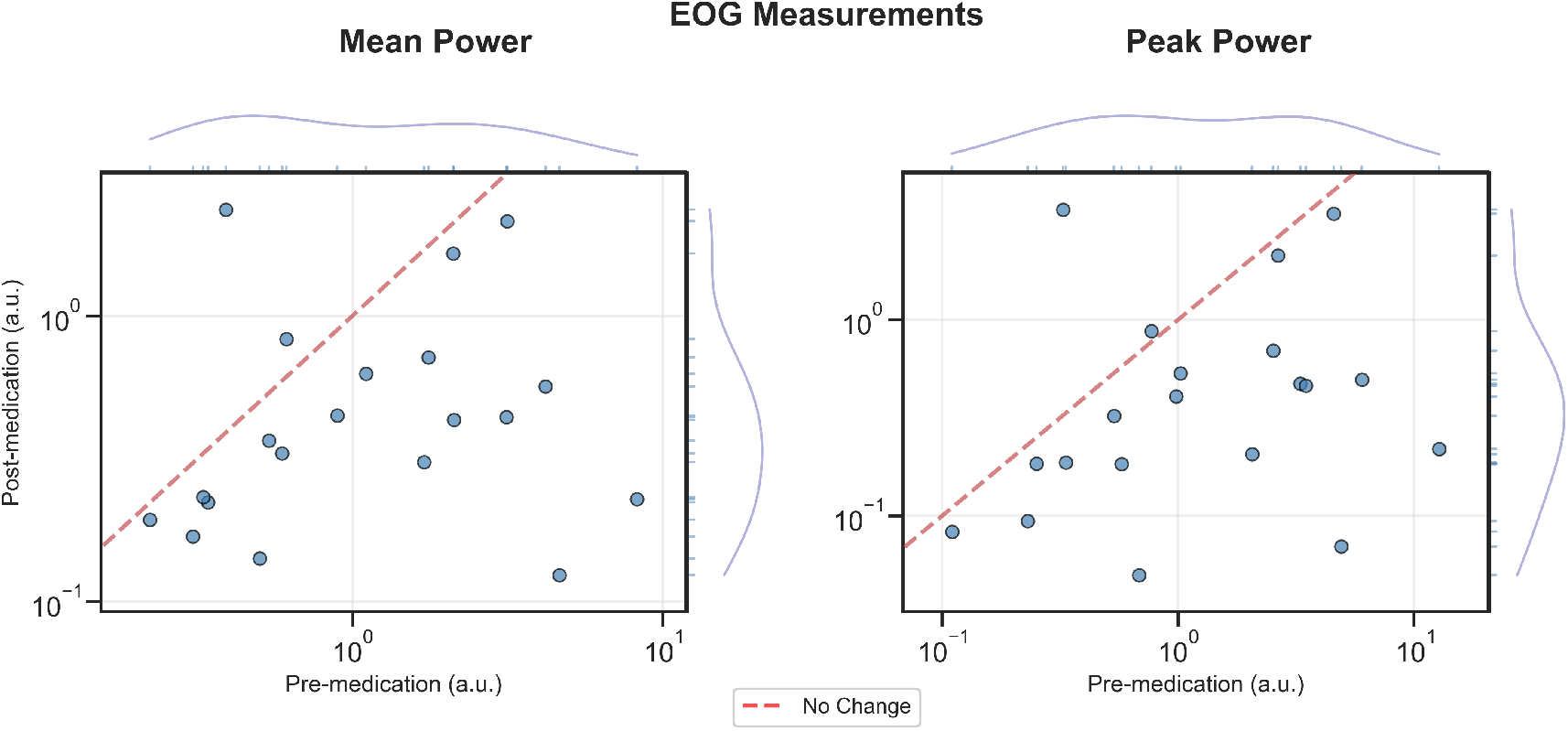
Pre- and Post-Medication EOG activity. Mean (left) and peak (right) EOG power before (horizontal axis) and after (vertical axis) medication for individual patients (n=20). Red dashed line represents no change. Marginal distributions display kernel density estimates with individual data points marked by rug plots. Log scale on both axes. Marginal distributions display kernel density estimates with individual data points marked by rug plots. Normalized values. Log scale on both axes. EOG: electrooculography; a.u.: arbitrary units.

### 3.3 Cardiovascular Changes

In contrast to the EOG findings, blood pressure showed no systematic change between sessions. This was true for both systolic pressure (t(19) = 0.188, p = 0.853; 13 increases, 7 decreases; binomial p = 0.263) and diastolic pressure (t(19) = 1.038, p = 0.312; 12 increases, 8 decreases; binomial p = 0.503).

Regarding heart rate, the mean pre–post difference was not statistically significant (t(19) = 1.651, p = 0.115). However, the binomial test indicated that significantly more participants showed an increase in heart rate (n = 15) than a decrease (n = 5), binomial p = 0.041. Taken together, these results suggest that although average changes were small and non-significant, a majority of participants exhibited an upward shift in heart rate between sessions.

### 3.4 EOG - Cardiovascular Correlation

In this control analysis, we examined whether changes in cardiovascular measures were associated with changes in the EOG parameters. No significant correlations were found between Δ pulse and either Δ mean EOG power (r = 0.059, p = 0.806, n = 20) or Δ peak EOG power (r = 0.025, p = 0.915, n = 20).

Similarly, the analyses involving blood pressure showed no systematic associations. Δ mean EOG power did not correlate significantly with Δ systolic pressure (r = 0.042, p = 0.860, n = 20) or Δ diastolic pressure (r = –0.148, p = 0.532, n = 20). Δ peak power showed equally negligible relationships with systolic (r = 0.043, p = 0.857, n = 20) and diastolic pressure (r = –0.179, p = 0.449, n = 20).

Overall, these results indicate that variability in the cardiovascular measures did not systematically relate to changes in the EOG parameters, as illustrated in **Figure 2**.

**Figure 2:**
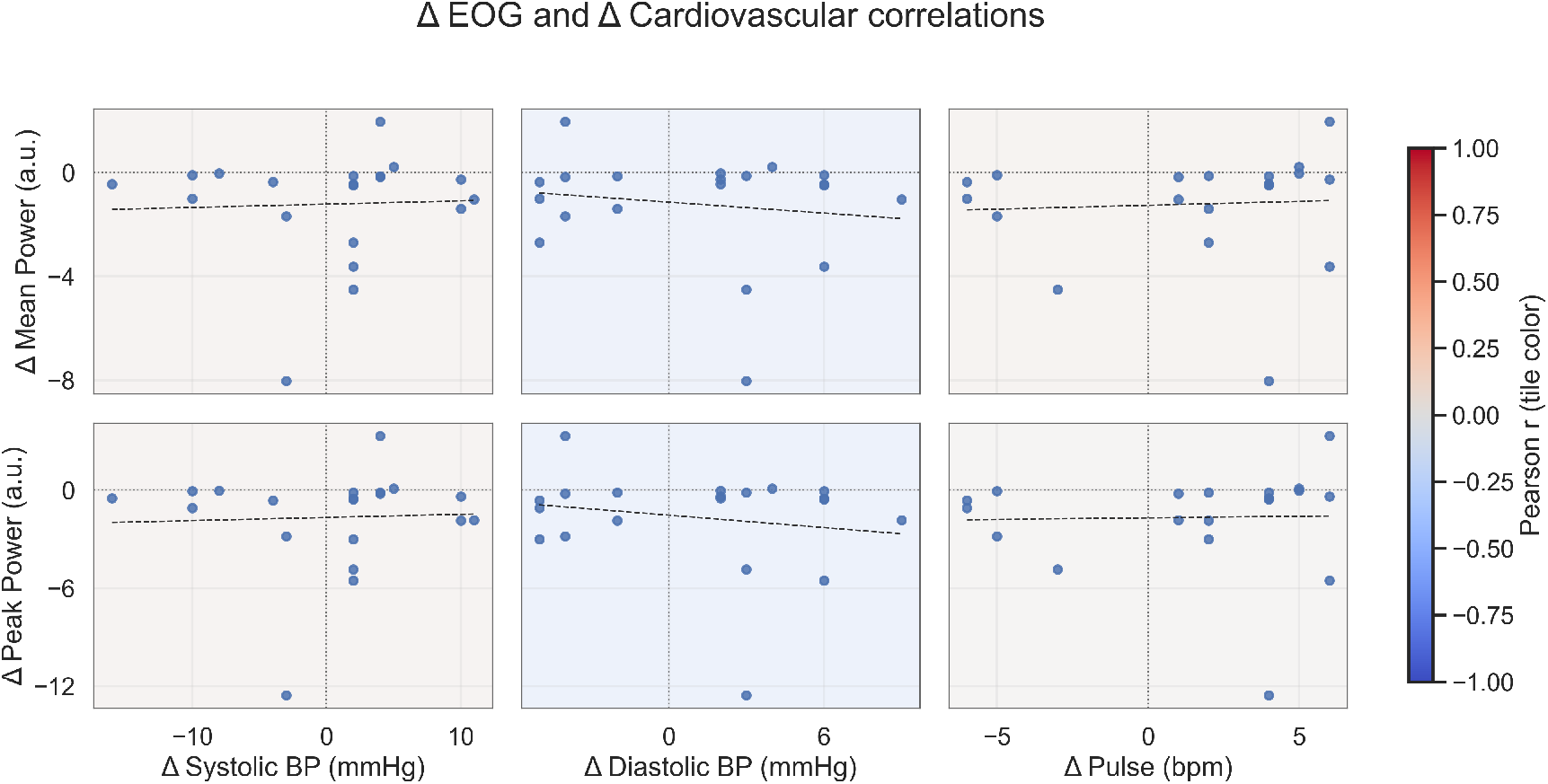
Correlation matrix of Δ EOG and Δ Cardiovascular parameters. Scatter plots display the relationship between changes in EOG measures (Δ mean power [top row] and Δ peak power [bottom row]) and cardiovascular parameters (Δ systolic blood pressure [left column], Δ diastolic blood pressure [middle column], and Δ pulse rate [right column]) between sessions (n=20). Each panel represents one parameter pair with individual subject data points (blue circles) and fitted linear regression lines (black dashed lines). Background tile colors indicate Pearson correlation coefficient strength (coolwarm colormap: blue = negative correlation, white = near-zero correlation, red = positive correlation). Horizontal and vertical dotted reference lines mark zero change. Normalized EOG values (a.u.). Linear scales on both axes. EOG: electrooculography; BP: blood pressure; Δ: change from pre-medication to post-medication; a.u.: arbitrary units.

### 3.5 Statistical Summary

Across participants, both mean and peak EOG power showed consistent and statistically significant reductions from pre-to post-medication, with 18 of 20 individuals exhibiting decreases in both measures. These findings indicate a robust within-subject change in EOG activity between sessions.

In contrast, cardiovascular parameters showed no systematic pre–post change. Systolic and diastolic blood pressure remained stable, and heart rate exhibited no significant mean-level difference despite a directional imbalance favouring increases. Supplementary control analyses further demonstrated that changes in cardiovascular measures did not correlate with changes in EOG parameters, indicating no detectable association between autonomic fluctuations and the observed EOG effects.

## 4. Discussion

### 4.1 Main Finding

This study demonstrates that EOG activity changes systematically following pharmacological treatment in patients with ADHD receiving stable, ongoing medication. By examining closed-eye, awake EOG recordings across a single clinical session, we show for the first time that this simple measurement setup can detect systematic within-subject changes over a short timeframe. The consistent reductions in EOG power observed across participants support our hypothesis that closed-eye EOG is sensitive to medication-related neurophysiological changes.

Cardiovascular parameters, in contrast, remained largely stable across sessions, and none of the cardiovascular changes correlated with the EOG measures. This indicates that the observed EOG modulation is unlikely to reflect general physiological arousal or autonomic fluctuations and instead may point to more specific neural effects of ADHD medication.

Together, these findings establish an initial link between pharmacological treatment and EOG dynamics in ADHD. They suggest that closed-eye EOG may offer a low-burden, non-invasive approach for detecting medication-related changes, providing groundwork for future studies to evaluate its clinical applicability and its relevance for individual treatment response patterns.

### 4.2 Interpretation For Titration

The observed within-session modulation of closed-eye EOG activity offers a potential conceptual link to pharmacological titration in ADHD. Because stimulant and non-stimulant medications alter neural circuits involved in arousal regulation, attention, and oculomotor control, a physiological measure that responds consistently within a short timeframe may reflect medication-related changes relevant to treatment adjustment. Closed-eye EOG is particularly suited to capturing such effects, as the absence of visual input or task demands minimises external influences and isolates intrinsic oculomotor dynamics that may be sensitive to pharmacological modulation.

From a titration perspective, the consistent direction and timing of the EOG changes suggest that this measure could indicate whether a given dose or medication produces a measurable neurophysiological effect. Importantly, the relative stability of cardiovascular parameters and the lack of association between cardiovascular and EOG changes suggest that the EOG modulation is not easily explained by general physiological arousal or other broad autonomic shifts. This strengthens the interpretation that closed-eye EOG may capture a more specific component of the medication response that is not directly reflected in peripheral physiological measures.

Taken together, these points indicate that closed-eye EOG could conceptually contribute additional information during medication titration by revealing short-latency neurophysiological shifts associated with treatment. While these findings do not establish clinical utility, they outline a rationale for further investigation into how EOG dynamics might complement behavioural and subjective assessments during dose optimisation in ADHD.

### 4.3 Comparison To Other Biomarkers

When viewed alongside other proposed biomarkers for ADHD, closed-eye EOG occupies a distinct position in terms of feasibility and potential clinical relevance. Genetic markers, such as single nucleotide polymorphisms (SNPs) in genes including DAT1 and DRD4, have been associated with differential responses to stimulant medication. However, their practical value is limited by modest effect sizes, population-level variability, and the need for specialised laboratory analyses, which restrict their immediate use in routine titration contexts [(8) (9)]

Neuroimaging approaches—particularly functional MRI and task-based or resting-state EEG—provide rich information about neural pathways implicated in ADHD and medication response. Yet their dependence on specialised equipment, technician expertise, and controlled environments poses substantial barriers to their integration into standard appointments [(29) (6)] Compared to these modalities, EOG requires minimal setup, can be recorded in an ordinary consultation room, and relies on straightforward analytical procedures, positioning it as substantially more accessible for clinical workflows.

Other proposed biomarkers, such as circulating microRNAs, remain at an exploratory stage. Although early findings suggest potential diagnostic or mechanistic relevance, the procedures involved—sample collection, laboratory processing, and assay interpretation—are resource-intensive and not yet suited for point-of-care use (30)

Taken together, existing biomarker candidates vary widely in promise, maturity, and feasibility. In this landscape, closed-eye EOG stands out not because it is more predictive or better validated—those questions remain open—but because it aligns well with practical constraints of ADHD care. Its simplicity, low burden, and compatibility with routine clinical settings make it a candidate biomarker with unusual logistical advantages. Future work will be needed to determine how its feasibility translates into clinical utility, particularly in comparison with more technically demanding but mechanistically richer approaches.

### 4.4 Limitations

A primary limitation of this study is the absence of a negative control or alternative control condition. Because all recordings took place within the natural flow of a clinical consultation, we cannot determine whether the observed EOG changes reflect a medication-specific effect or other factors inherent to the session, such as time, context, or expectancy. This constraint prevents causal inference and limits the strength of conclusions that can be drawn regarding specificity.

Relatedly, the present findings do not support any claims regarding prediction of individual medication response. Although systematic EOG changes were observed, the study design does not allow us to establish predictive validity, nor does it address whether the magnitude of EOG change relates to clinical improvement or symptom change. Any such interpretation would exceed the evidence provided here.

The cardiovascular measures included in the analysis served solely as contextual indicators and cannot be regarded as definitive markers of psychological or physiological states. Their variability across participants illustrates the complexity of autonomic processes in this setting and emphasises that the absence of correlation with EOG should not be taken as mechanistic evidence of independence.

Taken together, these limitations indicate that the findings should be interpreted cautiously and confined to the within-session physiological effects observed in this specific clinical context.

### 4.5 Future Directions

Future work should aim to clarify the clinical and scientific relevance of closed-eye EOG by examining its stability, specificity, and potential predictive value in more controlled settings. A key next step will be to conduct longitudinal studies to determine whether within-session EOG changes relate to subsequent clinical outcomes or longer-term patterns of treatment response. Such studies would provide insight into whether the short-latency effects observed here reflect transient physiological modulation or more enduring aspects of medication efficacy.

Larger cohorts, ideally across multiple sites, will also be important for assessing generalisability and identifying sources of inter-individual variability. These studies should incorporate appropriate control conditions or comparison groups to disentangle medication effects from session-related or contextual influences. In addition, integrating independent clinical outcome measures, such as symptom change, cognitive performance, or functional assessments, will be essential for determining whether EOG metrics contribute unique information beyond established clinical indicators.

Together, these steps will help establish whether closed-eye EOG can serve as a meaningful adjunct to existing methods of evaluating medication effects in ADHD and will inform the extent to which it may support more individualised treatment strategies in future work.

### 4.6 Conclusion

In conclusion, this study shows that closed-eye EOG activity changes systematically following medication intake in adults with ADHD who are receiving stable, ongoing treatment.

The consistent within-session reductions in EOG power across participants suggest that this simple physiological measure is sensitive to short-latency neurophysiological effects of medication. These findings support the idea that closed-eye EOG may capture medication-related changes in neural function in a manner compatible with routine clinical workflows.

The stability of cardiovascular measures, together with the absence of correlations between cardiovascular and EOG changes, indicates that the observed EOG modulation is unlikely to reflect general autonomic fluctuations. Instead, it may index a more specific aspect of treatment-related neural dynamics. While the present findings are preliminary, they provide a foundation for further investigation into the potential role of EOG in informing medication-related decision-making during clinical titration.

Establishing the clinical relevance of this approach will require controlled designs, longitudinal assessment, and larger, diverse cohorts. As this work progresses, closed-eye EOG may emerge as a low-burden, non-invasive adjunct to existing clinical methods for evaluating medication effects in ADHD, contributing to more nuanced and individualised treatment strategies in the future.

## 5. Statements

### 5.2 Ethics

This study was based exclusively on anonymised data derived from patient records. The original data were collected as part of standard clinical care for individuals treated for ADHD. As part of routine assessment and treatment, patients were informed about and given the option to undergo EOG recordings before and after medication, to be reviewed alongside other standard clinical measures such as body mass index (BMI), clinical rating scales, and medication data. The processing and anonymisation of patient record data were carried out as part of clinical quality assurance and development. Following anonymisation, all personal identifiers were removed, and the present study was conducted solely on these anonymised data. No identifiable information was available to the researchers, and no additional interventions or data collection were performed. In accordance with Danish regulations, studies based entirely on anonymised data do not require approval from a regional research ethics committee. The danish National Research Ethics Committee (National Videnskabsetisk Komité (NVK)) was consulted and confirmed that formal ethical approval was not required for this work.

### 5.4 Author Contribution

MB conducted data analysis, interpreted the results and produced the initial manuscript. SB performed the clinical and paraclinical assessments, maintained the anonymised clinical data base and generating the initial idea and concept for the study, Both authors reviewed and approved the final manuscript.

### 5.5 Data Availability

The data supporting the findings of this study are not publicly available, as they are part of an ongoing proprietary dataset under development for potential commercial use .

